# Same same but different: Variations in Therapists’ Application of the EMDR Standard Procedure and the Use of Minimal Nonverbal Changes

**DOI:** 10.64898/2026.01.06.26343513

**Authors:** Valeska Reichel Pape, Carolin von Klitzing, Olaf Wolkenhauer, Eva Schäflein, Antonia Hope, Markus Stingl

## Abstract

Following the standard Eye Movement Desensitization and Reprocessing (EMDR) procedure, counting a fixed number of bilateral stimuli (BLS) is the guideline-congruent strategy for determining the length of the first stimulation set during the processing of pathogenic memories. For the subsequent sets, an individually adapted stimulation length depending on minimal nonverbal changes (MNCs) is recommended, such as changes in facial expression, muscle tone, posture, or breathing, which are assumed to reflect shifts in emotional valence. This exploratory survey study is the first work to investigate the relevance of these MNCs in clinical practice.

This exploratory survey study examined how certified EMDR therapists apply the BLS procedure in practice and how they evaluate the relevance of MNCs. Eighty-eight certified EMDR therapists reported on their individual application of BLS. In addition, the perceived frequency, indicator value, and emotional valence of a broad range of MNCs were assessed.

The results revealed substantial variability in therapists’ application of the BLS procedure. Whereas one quarter of therapists reported using counting as their preferred interruption strategy, approximately three quarters relied on MNCs. Nearly all therapists (98.2%) considered MNCs clinically important. Forehead relaxation, smiling, and respiratory changes were rated as particularly relevant indicators of valence change. Exploratory analyses suggested that therapist gender, therapeutic orientation, and patient clientele may be associated with variation in interruption strategies.

These findings provide initial descriptive evidence of substantial heterogeneity in how EMDR therapists determine the length of BLS sets in clinical practice. They also identify specific MNCs that may be relevant targets for future observational and experimental research.

## INTRODUCTION

In recent years, several survey studies have examined the use of Eye Movement Desensitization and Reprocessing (EMDR) [1-4] from the perspective of practicing therapists. These studies have explored therapists’ experiences in specific contexts, such as climate-related anxiety [5], particular applications including online EMDR [6] or couple therapy [7], and work with specific patient populations, such as individuals with autism [8] or intellectual disabilities [9].

Other surveys have focused on factors potentially associated with treatment outcomes, including the quality of the therapeutic relationship [10], supervision and training [11], and characteristics of the clinical setting and therapeutic orientation [12]. For example, analytically or humanistically oriented therapists have been reported to experience more difficulties than cognitive-behavioral clinicians, while therapists in private practice appear more engaged in EMDR-related activities than those working in institutional settings [12].

However, little is known about how therapists implement the EMDR standard procedure in routine clinical practice. In particular, there is a lack of systematic data on individual differences in the application of bilateral stimulation (BLS), including the extent to which clinical practice aligns with procedure recommendations. As a result, it remains unclear how variations in procedure implementation may relate to therapeutic processes and outcomes.

The standard EMDR procedure as described by Shapiro [3] constitutes the foundation of EMDR therapy for patients with posttraumatic stress disorder (PTSD) and serves as the basis for various adaptations, for example in highly dissociative patients [13-15], anxiety disorders [16], and addictive disorders [17-18]. A core element of EMDR is the application of BLS during imagery-based exposure, which can be delivered visually, tactilely, or auditorily. One aspect of the procedure that allows for considerable individual interpretation is the determination of the optimal duration of each stimulation set. The standard procedure recommends two strategies: a) counting a fixed number of BLS (e.g., approximately 24 stimuli) in the initial set, b) followed by a variable number in subsequent sets, while explicitly emphasizing that these parameters should be adapted to the ongoing emotional processing (i.e. emotional valence) of the patient, indicated by minimal nonverbal changes (MNCs) [3]. In this approach, therapists adjust the duration of BLS sets dynamically based on observable changes in facial expression, breathing, posture, or muscle tension. Findings from emotion research suggest that such nonverbal cues can reflect shifts in emotional valence [19–21], and may therefore provide clinically relevant information for timing the interruption of stimulation. However, empirical evidence supporting the relevance of MNCs during EMDR therapy remains limited. In particular, it remains unclear how frequently therapists rely on counting versus MNC-guided strategies, and to what extent individual variations in implementation align with existing guidelines.

### Aims

The present survey pursued two main objectives:

#### Objective 1

To describe how EMDR therapists implement the standard procedure in routine clinical practice, with a particular focus on the application of BLS. In addition, potential sources of variability were explored, including therapist characteristics, clinical setting, and patient population. The aim was to provide a descriptive account of current practice patterns and to identify factors that may contribute to individual differences in procedure implementation. This objective is exploratory in nature and intended to inform the development of hypotheses for subsequent, confirmatory studies.

#### Objective 2

To obtain initial data on the clinical relevance of MNCs within EMDR therapy. The goal was to generate a preliminary ranking of MNCs and to explore their potential role as indicators of ongoing emotional processing within EMDR therapy. Based on prior findings from emotion research [19-21], theory-informed expectations were formulated. Specifically, relaxation of the face, smiling, deep exhalation/ inhalation, or relaxation of the body were expected to be associated with positive valence, whereas facial distortion, increase in muscle tone, or increased respiratory rate were expected to be associated with negative valence. Given the exploratory design of the study, these hypotheses were not tested in a confirmatory manner but served as a conceptual framework for interpreting the descriptive findings.

## MATERIALS AND METHODS

### Study design

The overall study design is illustrated in Figure 1.

**Fig. 1.**
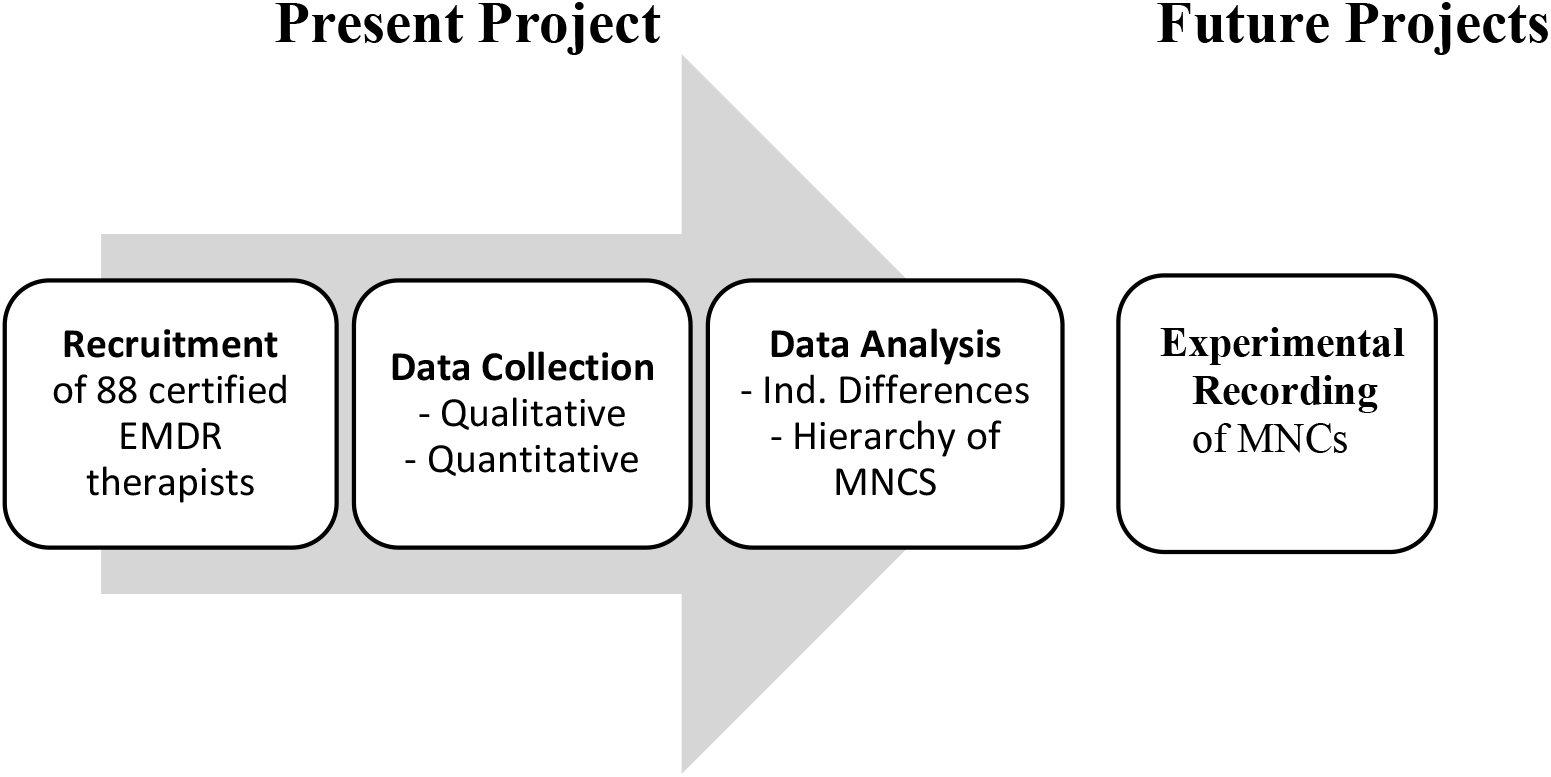
Experimental and technical design. The figure illustrates the design of the present study and provides a preview of potential future projects (see Discussion).

#### Questionnaire development

A study-specific, two-part questionnaire was developed using an iterative, expert-informed process in line with established survey development principles [22-24]. An initial item pool was generated by the research team to assess acceptability of and individual adaptations to the EMDR standard procedure (qualitative component) and evaluate the reported frequency, perceived indicator value, and emotional valence of MNCs (quantitative component), alongside basic therapist characteristics.

The draft questionnaire was reviewed by an expert panel consisting of four EMDR practitioners and one EMDR supervisor, all with several years of clinical experience. The panel provided qualitative feedback on clinical relevance, content coverage, wording, and response options, and suggested additional items. Based on this feedback, the questionnaire was revised in two iterative rounds to improve clarity and reduce redundancy before being implemented as an online survey.

No formal pilot testing or quantitative item validation was conducted.

### Sample characteristics

A total of 88 EMDR therapists (aged 32-74 years) from German-speaking countries were included in the study. Participants were recruited via publicly accessible therapist directories of professional EMDR associations in Germany, Switzerland, and Austria. Inclusion criteria were certification in EMDR by a recognized training institute and current clinical practice in trauma-focused therapy. Therapists without formal EMDR certification were excluded. All participants provided written informed consent. Ethical approval was obtained from the ethics committee of the University Medical Center Rostock. Detailed sample characteristics are presented in Table 1.

**Table 1.** Sample characteristics. Values are presented as frequencies (percentages) or means (*M*) +/-standard deviations (*SD*). PT = psychotherapeutic. PTSD = posttraumatic stress disorder.

| Variable | Value |
| --- | --- |
| Age (years) | 54.6 Years +/- 10.1 Years |
| Gender (%) | Female (73 %), Male (25.8 %), Non-Binary (1.2 %) |
| Location (%) | Northern Germany (65.2 %), Southern Germany (29.3 %), Austria (1.7 %), Switzerland (1.7 %) |
| Training (%) | EMDR Institute (70.8 %), Institute of for Trauma Therapy (4.5 %), other institutes (23.6 %) |
| Education (%) | Psychologists (56.2 %), Physicians (41.6 %) |
| PT Orientation (%) | Psychoanalytic (24.7 %), Behavioral (52.8 %) |
| Training Time (years) | 11.6 Years +/- 7.1 Years |
| Patient Clientele (%) | Only Single Trauma (4 %), also complex PTSD (95.5 %) |
| Setting (%) | Inpatients (87.5 %), Outpatients (12.5 %) |

### Survey methods

The survey instrument (*EmdEx*) was developed to capture therapists’ experiences with and application of the EMDR standard procedure, as well as their evaluation of MNCs. The questionnaire included both qualitative and quantitative components and was designed to allow a structured assessment of practice patterns and perceived indicators of emotional processing.

#### Demographic and professional characteristics

At the beginning of the survey, participants provided information on demographic and professional characteristics. These variables were assessed using multiple-choice items with nominal or ordinal response formats.

- Demographic data: age, gender, and country of practice (Germany, Switzerland, or Austria)
- Professional background: EMDR training institute, academic qualification (physician or psychologist), therapeutic orientation (behavioral or psychodynamic), and years of experience in the treatment of PTSD
- Patient population: primary patient groups treated (‘Type 1 PTSD’, ‘Type 2 PTSD’, ‘complex PTSD’, ‘other’, or a combination)
- Practice setting: type of clinical setting (‘private practice’, ‘outpatient clinic’, ‘inpatient clinic’, or a combination)

#### Qualitative component

In the qualitative component, therapists’ individual application and evaluation of the EMDR standard protocol were assessed, with a focus on interruption strategies during BLS and the use of MNCs.

- Acceptability of procedure-based interruption strategies: Therapists indicated whether they were satisfied with the standard procedure recommendations for interrupting BLS or required more precise criteria. They were also asked whether they would consider biofeedback as an objective alternative. Participants who did not endorse biofeedback were asked to specify their reasons (‘would irritate me’/’would irritate the patient’). All items were assessed using dichotomous (yes/no) response formats.
- Perceived consequences of suboptimal timing: Therapists reported whether they considered interrupting BLS too early or too late to be problematic and, if so, which consequences they expected. Several predefined consequences were assessed: prolonged session duration, missed valence shifts, insufficient reprocessing or looping, insufficient decrease in Subjective Units of Distress (SUD), insufficient increase in Validity of Cognition (VoC), or no consequences.
- Interruption strategies: Participants selected their preferred strategy for determining the duration of BLS sets (multiple-choice): counting a fixed number of stimuli, orienting to MNCs, orienting to internal judgment, or a combination of these approaches. For those using counting strategies, the specific number of BLS was recorded.
- Perceived relevance of MNCs: Therapists indicated the relevance of MNCs (no relevance, indication of increased distress, indication of valence change, or a combination).
- Predictive value of MNCs: Participants rated the temporal relationship between MNCs and emotional responses (occurring before, after, or simultaneously with the emotional reaction, combinations thereof, or no predictive value).
- Estimated latency: Therapists estimated the time interval (in seconds) between emotional responses and the occurrence of MNCs.
- Handling of MNCs: Finally, therapists reported how they typically respond to observed MNCs (ignore, continue stimulation, interrupt stimulation, or other). If ‘interruption’ was selected, the number of BLS between observing an MNC and interrupting stimulation was recorded.

#### Quantitative component

In the quantitative component, therapists evaluated a predefined list of MNCs with regard to their frequency, perceived indicator value, and associated emotional valence. The list was derived from findings in emotion research suggesting that emotional processes can be inferred from nonverbal behavior [19-21]. Participants were also given the opportunity to add additional MNCs not included in the predefined list.

The MNCs covered the following domains:

- Facial expressions: facial tension, facial relaxation, smiling, mouth distortion, crying, frowning, forehead relaxation, puffing of the cheeks, eye closure, eye widening, increased blink rate, decreased blink rate
- Breathing: deep inhalation, deep exhalation, increased respiratory rate (hyperventilation), decreased respiratory rate (hypoventilation), breath holding
- Gestures: opening or clenching fists, self-touch (face or body), flexion or extension of the arms or legs, finger or foot tapping, crossing the legs, parallel leg positioning, head tilt toward or away from the therapist, nodding, head shaking
- Posture: changes in sitting position, freezing, upright or slouched posture, sudden loss or increase of muscle tone, forward or backward shifting, trembling or flinching, body orientation toward or away from the therapist, evasive movements
- Autonomic responses: visible sweating, freezing reactions
- Vocalizations: laughing, moaning, sobbing, sighing

For each MNC, three ratings were obtained:

- Frequency: rated on a scale from 0 (never) to 10 (very often)
- Indicator value: perceived ability to reflect changes in emotional valence, rated from 0 (not suitable at all) to 10 (perfectly suitable)
- Emotional valence: categorized as indicating a negative, neutral, or positive emotional response

### Statistical analysis

#### Descriptive statistics

Data were analyzed using SPSS (IBM Corp., Armonk, NY, USA). Descriptive statistics were used to summarize sample characteristics and questionnaire responses. Categorical variables are reported as frequencies and percentages. Rating-scale data (e.g., frequency and indicator values of MNCs) were summarized using mean values for graphical representation [25-27].

#### Qualitative component

Exploratory inferential analyses were conducted to examine potential associations between therapist characteristics (e.g., age, gender, therapeutic orientation, clinical setting, and patient population) and selected questionnaire responses from the qualitative component. For continuous variables (e.g., number of BLS, estimated latency in seconds), univariate analyses of variance (ANOVA) and correlation analyses were applied, with Partial Eta-squared (*η*^*2*^*p*) as the primary effect size measure. For categorical variables, non-parametric tests (*χ*^*2*^ tests) were used based on contingency tables, with Cramer’s V (*V*, for larger tables) and Phi coefficient (*φ*, for 2 x 2 tables) as the primary effect size measures. For proportional data, Cohen’s h (*h*) was calculated (*p*_*1*_ = proportion 1; *p*_*2*_ = proportion 2). Given the exploratory and hypothesis-generating nature of this component of the study, no correction for multiple testing was applied [28-31]. Accordingly, reported p-values should be interpreted descriptively and with caution.

#### Quantitative component

Frequency and indicator values of MNCs were plotted on a two-dimensional graph (frequency on the x-axis, indicator value on the y-axis). MNCs with a frequency > 6 were classified as frequent, and those with an indicator value > 6 as clinically relevant. Based on these criteria, a rank order of MNCs was established, stratified by emotional valence (positive vs. negative), and the Wilcoxon signed-rank test was performed to compare MNCswith high ranks vs. low ranks. As correction for multiple testing, Bonferroni-corrections were conducted here.

## RESULTS

### Qualitative component

#### Descriptive findings

Acceptability of procedure-based interruption strategies: Only 21.8% (*p*_*1*_ = 0.22) of therapists expressed a need for a more explicit criterion for terminating BLS, whereas 78.1% (*p*_*2*_ = 0.78) reported no such need (74.7%) or were unsure (3.4 %). The magnitude of this effect (*h* = 1.188) indicates a high acceptance of procedure-based interruption strategies. Regarding the use of biofeedback or other objective indicators of valence change, responses were mixed: 42.0 % (*p*_*1*_ = 0.42) expressed interest, and 58% (*p*_*2*_ = 0.58) expressed no interest (48.9%) or were undecided (9.1%) (Figure 2). This represents a small effect size (*h* = 0.321), indicating a high ambivalence under EMDR therapists, with the one half being interested in biofeedback and the other half being not. Among those not endorsing biofeedback, the most frequently reported reasons were perceived interference with therapeutic work (28.4 %), potential irritation of the patient (4.5 %), or both therapist and patient (20.5 %). The remaining 46.6% did not provide a specific reason.

**Fig. 2:**
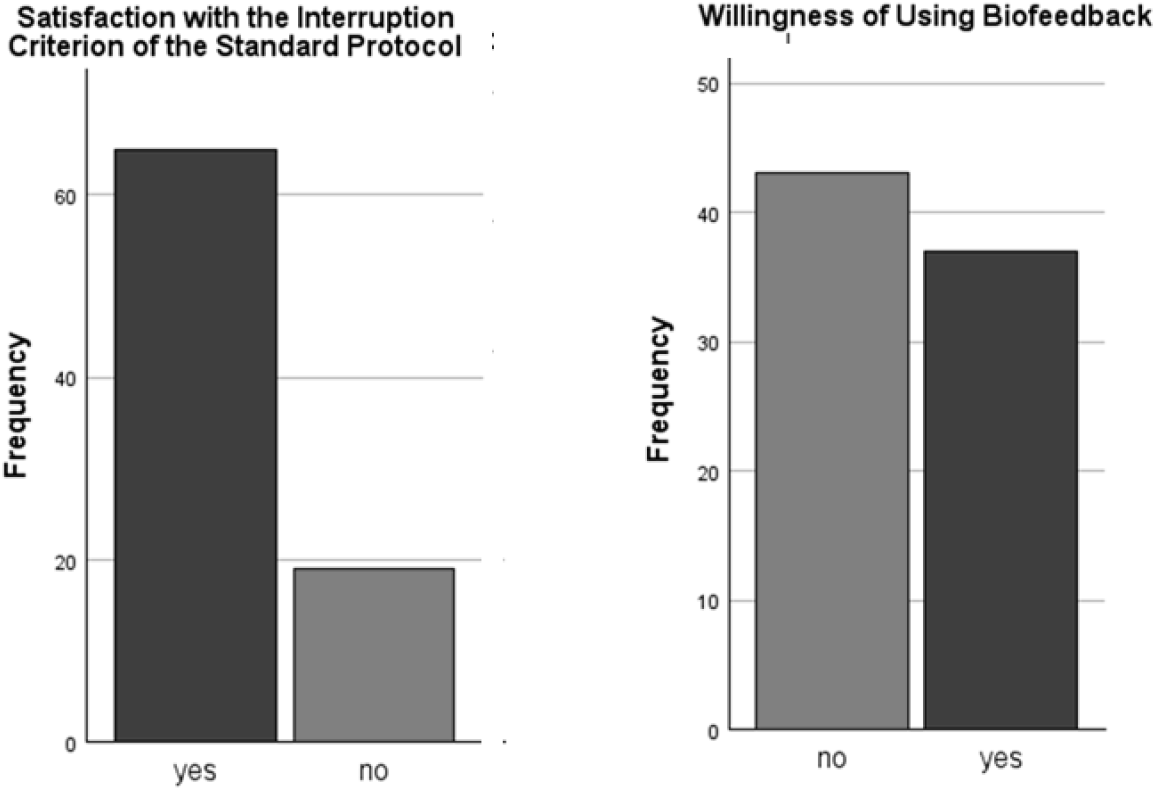
Acceptability of the standard protocol and willingness to use biofeedback. Values are presented as frequencies. Dichotomous (yes/no) questions were used. Therapists were asked whether they were satisfied with the standard protocol, and whether they would be interested in using biofeedback as an objective alternative.

Perceived consequences of suboptimal timing: A large majority of therapists (90.1%, *p*_*1*_ = 0.90) considered premature or delayed interruption of BLS to be problematic and potentially detrimental to treatment, whereas 9.2% (*p*_*2*_ = 0.09) did not. The resulting effect size (*h* = 1.888) far exceeds Cohen’s threshold for a large effect, indicating an exceptionally strong difference between proportions and suggesting near-universal awareness of the negative consequences associated with suboptimal timing. Reported consequences included insufficient reprocessing or looping (83.0%), prolonged stimulation (59.1%), missed valence changes (51.1%), and insufficient reduction in SUD (45.5%).

Interruption strategies: 45.5 % of therapists reported using MNC-based approaches as their preferred strategy for determining when to interrupt BLS sets. Only 13.6 % of therapists reported using counting-based approaches as their preferred strategy. 51.1% reported using a combination of both approaches, and a small proportion (3.4%) indicated reliance on intuitive or situational judgment. The comparison between using MNCs (*p*_*1*_ = 0.46) vs. counting down (*p*_*2*_ = 0.14) indicates a large effect (*h* = 0.80) with strong preference for MNCs.

Perceived relevance of MNCs: The vast majority of therapists (982%, *p*_*1*_ = 0.982) rated MNCs as highly important in their clinical practice, whereas 1.1% did not (*p*_*2*_ = 0.018) (Figure 3). This represents an exceptionally large effect size (*h* = 0.80), indicating a very strong consensus among EMDR therapists regarding the clinical relevance of nonverbal indicators. Most respondents (91.0%) indicated that MNCs may reflect both ongoing negative valence and shifts toward positive valence, while 6.7% associated MNCs exclusively with positive valence.

**Fig. 3:**
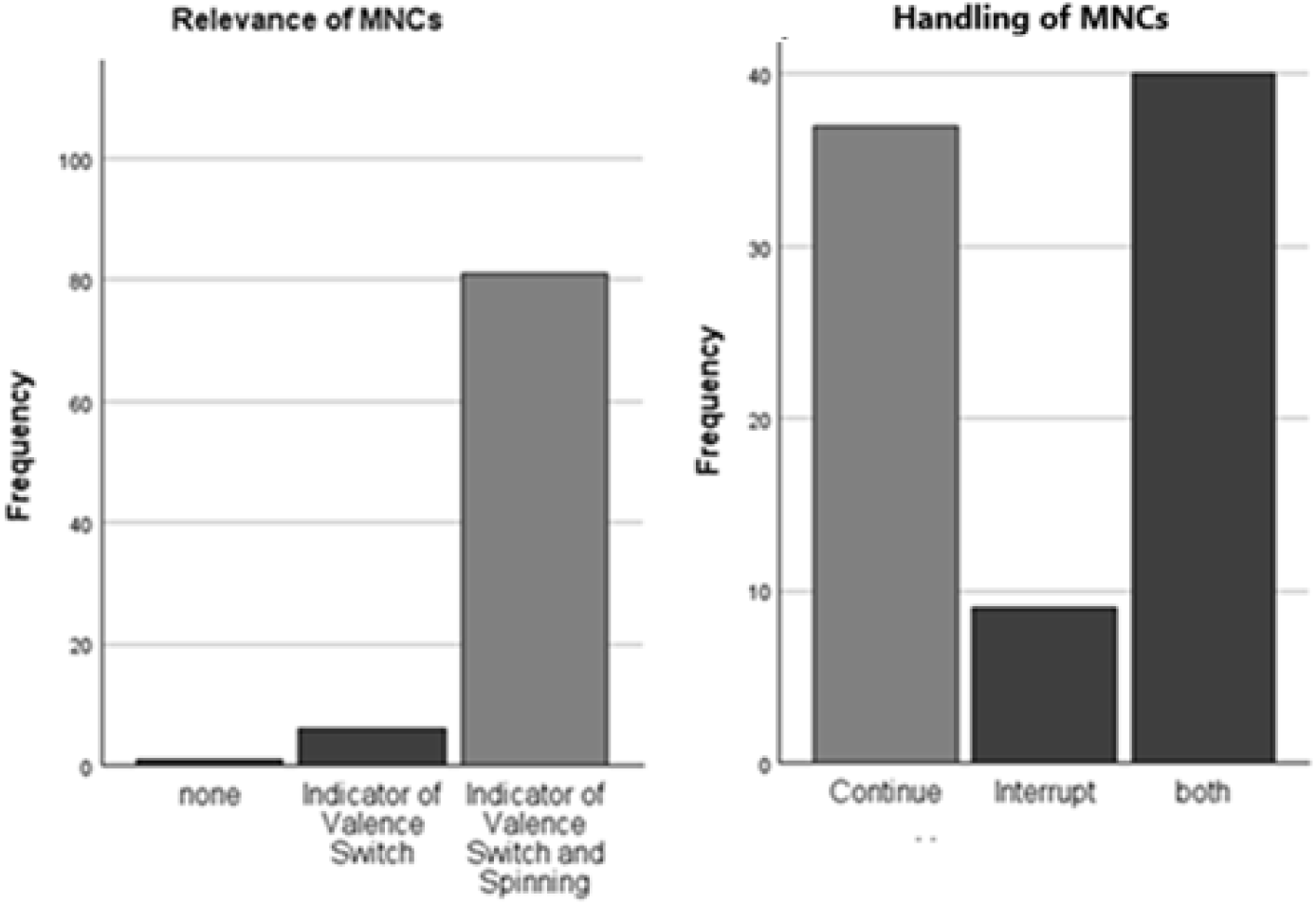
Perceived relevance of and responses to minimal nonverbal changes (MNCs) Perceived relevance of and responses minimal nonverbal changes (MNCs). Values are presented as frequencies. Multiple-choice questions were used. Therapists were asked to indicate the relevance they attributed to MNCs, and how they respond after observing an MNC.

Handling of MNCs: Therapists reported heterogeneous strategies in response to observed MNCs. A total of 51.7% (*p*_*1*_ = 0.52) reported either interrupting (10.1%,) or continuing (41.6%) stimulation upon observing an MNC, whereas 44.9% (*p*_*2*_ = 0.45) indicated flexible handling depending on the clinical situation. The effect size for this analysis (*h* = .140) was found to be below the range of Cohen’s convention for a small effect, indicating a high variability in the handling of MNCs.

Predictive value and temporal dynamics of MNCs: Therapists estimated that MNCs may occur both before and after changes in emotional valence, with an approximate window of 7.5 seconds before and 10 seconds after the valence shift. The estimated latency between emotional change and the occurrence of an MNC was on average 3.2 seconds (*SD* = 2.5 seconds; range: 0.5-10 seconds).

#### Exploratory inferential analysis

The findings of the inferential analysis are depicted in Table 2.

**Table 2.**
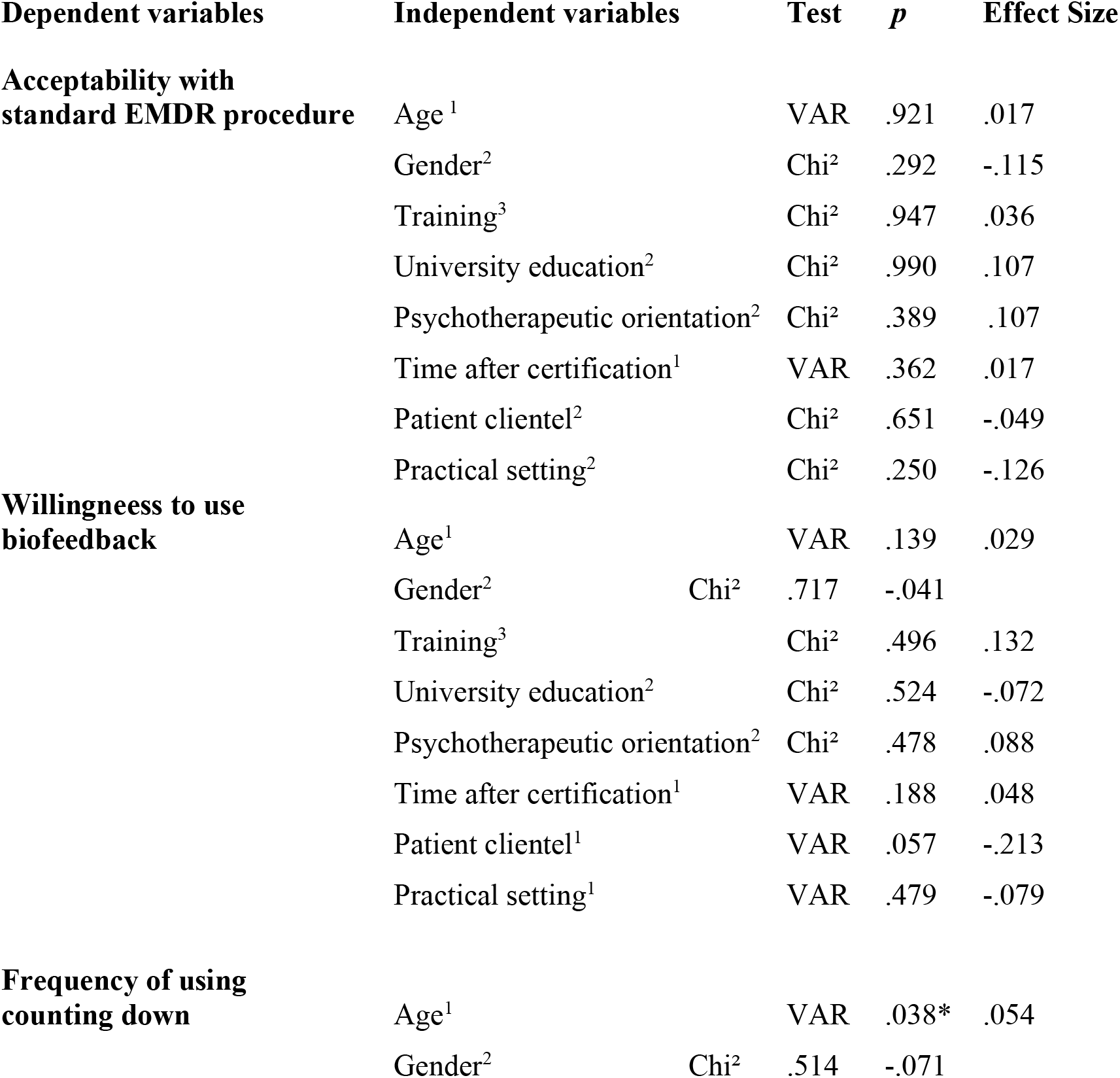
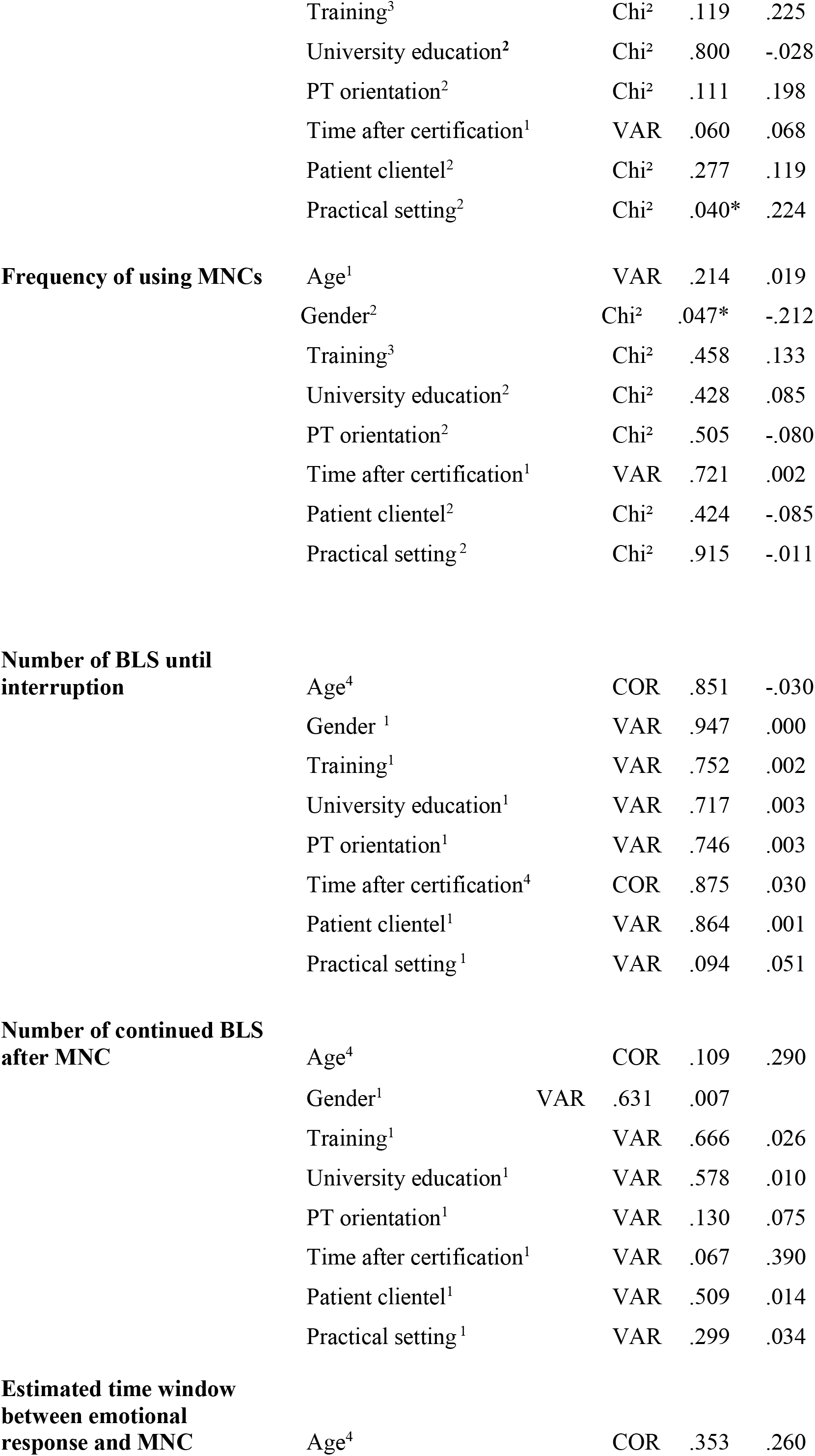
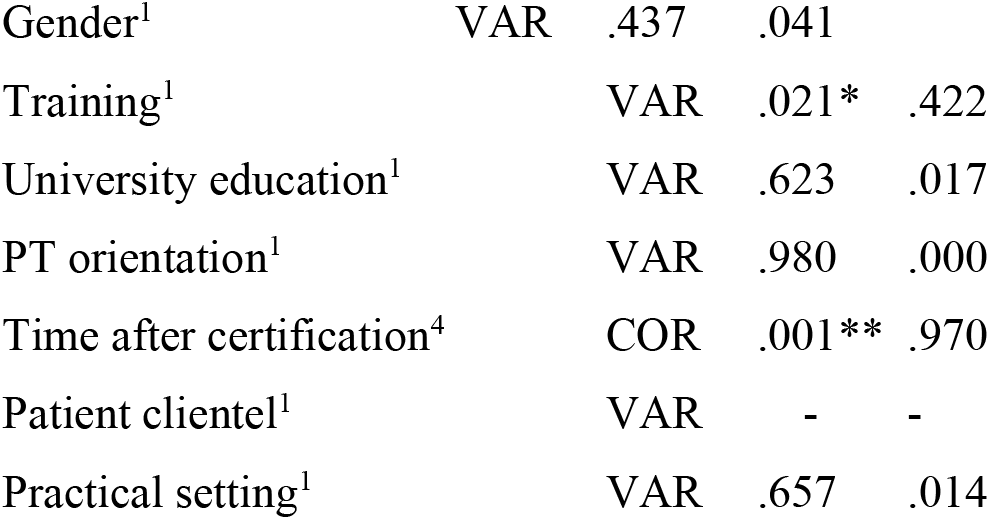
Results of inferential-statistical analyses. Results of the explorative inferential statistical analysis. The table illustrates associations between therapist characteristics (sociodemographic variables, training background, university education, psychotherapeutic orientation), patient characteristics, and treatment setting with several dependent variables, including acceptability of standard procedure-conform interruption strategies, preferred interruption strategies, handling of minimal nonverbal changes (MNC), and estimated time windows between emotional response and MNC. Analyses were conducted using via univariate analyses of variance (VAR), correlation analyses (COR), and chi^2^-tests (Chi^2^). Effect sizes were calculated via: ^1^Partial Eta Square (*η*^*2*^*p*), ^2^Phi (*φ*), ^3^Cramer’s V (*V*), or Correlation Coefficent^4^ (*r*). * *p* < .050, ** *p* < .001

Age:Therapist age was not associated with acceptability of procedure-based interruption strategies (*F*[1,58] < 1, *p* = .921, *η*^*2*^*p* = .017), willingness to use biofeedback (*F*[1,58] < 1, *p* = .139, *η*^*2*^*p* = .029), or the reported use of MNCs (*F*[1,58] = 1.13, *p* = .214, *η*^*2*^*p* = 019) and counting strategies (*F*[1,58] = 4.46 *p* = .038, *η*^*2*^*p* = .054). No correlations were found between age and the number of BLS applied before interruption (*r* = -0.03, *p* = .851), the estimated latency between emotional response and MNC (*r* = .26, *p* = .353), or the number of additional BLS administered after observing an MNC (*r* = .29, *p* = .109).

Gender: Gender was not associated with acceptability of procedure-based interruption strategies (*χ*^*2*^[1] = 1.11, *p* = .292, *φ* = -.115), willingness to use biofeedback (χ^2^[1] = 0.13, *p* = .717, *φ* = -.041), or the use of counting strategies (*χ*^*2*^[1] = 0.43, *p* = .514, *φ* = -.071). An exploratory association was observed between gender and the use of MNCs, with female therapists reporting more frequent use MNCs than male therapists (*χ*^*2*^[1] = 3.95, *p* = .047, *φ* = .212, i.e. small effect). No associations were found between gender and the number of BLS applied before interruption (*F*[1,54] < 1, *p* = .947, *η*^*2*^*p* = .000), the number of additional BLS administered after observing an MNC (*F*[1,32] < 1, *p* = .631, *η*^*2*^*p* = .007), or the estimated latency between emotional response and MNC (*F*[1,15] < 1, *p* = .437, *η*^*2*^*p* = .041).

Training institute: The EMDR training institute was not associated with acceptability of procedure-based interruption strategies (*χ*^*2*^[2] = 0.11, *p* = .947, *V* = .036), willingness to use biofeedback (*χ*^*2*^[2] = 1.40, *p* = .496, *V* = .132), or the use of counting strategies (*χ*^*2*^[4] = 1.30, *p* = .119, *V* = .225) or MNCs (*χ*^*2*^[2] = 1.56, *p* = .458, *V* = .133). No associations were found with the number of BLS applied before interruption (*F*[1,56] < 1, *p* = .752, *η*^*2*^*p* = .002) or the number of additional BLS administered after observing an MNC (*F*[2,31] < 1, *p* = .666, *η*^*2*^*p* = .026). An exploratory association was observed for the estimated latency between emotional response and MNCs (*F*[2,14] = 5.12, *p* = .021, *η*^*2*^*p* = .422, i.e. medium effect), with therapists trained at the Institute of Trauma Therapy Berlin reporting longer latency estimates than those trained at other institutes.

Professional background (physician vs. psychologist): Professional background was not associated with the use of counting strategies (*χ*^*2*^[1] = 0.06, *p* = .990, *φ* = .107) or MNCs (*χ*^*2*^[1] = 0.63, *p* = .428, *φ* = .085), acceptability of procedure-based interruption strategies (*χ*^*2*^[1] < 0.01, *p* = .990, *φ* = .107), or willingness to use biofeedback (*χ*^*2*^[1] = 0.41, *p* = .524, *φ* = -.072). No associations were found with the number of BLS applied before interruption (*F*[1,53] < 1, *p* = .717, *η*^*2*^*p* = .003), the number of additional BLS after observing an MNC (*F*[1,32] < 1, *p* = .578, *η*^*2*^*p* = .010), or the estimated latency between emotional response and MNC (*F*[1,15] < 1, *p* = .623, *η*^*2*^*p* = .017).

Therapeutic orientation: Therapeutic orientation was not associated with acceptability of procedure-based interruption strategies (*χ*^*2*^[1] = 0.74, *p* = .389, *φ* = .107), willingness to use biofeedback (*χ*^*2*^[1] = 0.50, *p* = .478, *φ* = .088), or the use of MNCs (*χ*^*2*^[1] = 0.45, *p* = .505, *φ* = -.080). An exploratory trend was observed for the use of counting strategies (*χ*^*2*^[1] = 2.54, *p* = .111, *φ* = .189) with behavioral therapists reporting more frequent use than psychodynamic therapists. Descriptively, psychodynamic therapists reported equal use of counting and MNC-based strategies, whereas behavioral therapists more frequently relied on counting strategies. No associations were found with the number of BLS applied before interruption (*F*[1,42] < 1, *p* = .746, *η*^*2*^*p* = .003), the number of additional BLS after observing an MNC (*F*[1,30] = 4.42, *p* = .130, *η*^*2*^*p* = .075), or the estimated latency between emotional response and MNC (*F*[1,12] < 1, *p* = .980, *η*^*2*^*p* = .000).

Time since certification: Time since certification was not associated with acceptability of procedure-based interruption strategies (*F*[1,33] < 1, *p* = .362, *η*^*2*^*p* = .017), willingness to use biofeedback (*F*[1,33] < 1, *p* = .188, *η*^*2*^*p* = .048), or the use of MNCs (*F*[1,33] < 1, *p* = .721, *η*^*2*^*p* = .002) and counting strategies (*F*[1,33] = 2.85, *p* = .060, *η*^*2*^*p* = .068). A strong correlation was observed between time since certification and estimated latency between emotional response and MNCs (*r* = .970, *p* = .001). No correlations were found with the number of BLS applied before interruption (*r* = .030, p = .875) or the number of additional BLS after observing an MNC (*r* = .390, *p* = .067).

Patient population: Patient population was not associated with acceptability of procedure-based interruption strategies (*χ*^*2*^[1] = 0.20, *p* = .651, *φ* = -.049), or the use of counting strategies (*χ*^*2*^[1] = 1.18, *p* = .277, *φ* = .119) or MNCs (*χ*^*2*^[1] = 0.64, *p* = .424, *φ* = -.084). An exploratory trend was observed for willingness to use biofeedback (*χ*^*2*^[1] = 3.62, *p* = .057, φ = .213), with therapists treating single-event trauma more likely to endorse biofeedback than those treating PTSD. No associations were found with the number of BLS applied before interruption (*F*[1,54] < 1, *p* = .864, *η*^*2*^*p* = .001) or the number of additional BLS after observing an MNC (*F*[1,32] < 1, *p* = .509, *η*^*2*^*p* = .014). The relationship between patient population and estimated latency could not be analyzed due to insufficient valid cases.

Clinical setting: Clinical setting (inpatient vs. outpatient) was not associated with acceptability of procedure-based interruption strategies (*χ*^*2*^[1] = 1.32, *p* = .250, *φ* = -.126), willingness to use biofeedback (*χ*^*2*^[1] = 0.50, *p* = .479, *φ* = -.079), or the use of MNCs (*χ*^*2*^[1] = 0.01, *p* = .915, *φ* = -.011). An exploratory association was observed for the use of counting strategies (*χ*^*2*^[1] = 4.21, *p* = .040, *φ* = .224), with therapists in inpatient settings reporting more frequent use of counting. No associations were found with the number of BLS applied before interruption (*F*[1,25] < 1, *p* = .094, *η*^*2*^*p* = 051), the number of additional BLS after observing an MNC (*F*[1,32] = 1.11, *p* = .299, *η*^*2*^*p* = .034), or the estimated latency between emotional response and MNC (*F*[1,15] < 1, *p* = .657, *η*^*2*^*p* = 014).

### Quantitative component

MNCs were ranked based on therapists’ ratings of frequency, indicator value, and associated emotional valence. In addition, potential physiological correlates were descriptively assigned where applicable. The resulting rank order is depicted in Figure 4 and Table 3. Separately for each variable, the data was divided in MNCs with high ranks vs. low ranks, and non-parametric tests were performed, corrected for multiple measurements. The findings indicate a reliable separation between MNCs with high vs. low frequency (*z* = 4.428, *p* < .001), high vs. low indicator value (*z* = 4.474, *p* < .001), and positive vs. negative valence (*z* = 4.899, *p* < .001).

**Table 3.** Rank order of minimal nonverbal changes (MNCs). The ranking is based on the estimated frequency and perceived indicator value of each MNC, calculated separately for each emotional valence (positive vs. negative). MNCs with higher frequency and a higher indicator value within each valence category are ranked higher.

| Rank | Positive Valence | Negative Valence |
| --- | --- | --- |
| 1 | Relaxation of the Face | Facial Distortion |
| 2 | Smiling | Sudden Increase in Muscle Tone |
| 3 | Deep Exhalation | Sobbing |
| 4 | Deep Inhalation | Increasing Respiratory Rate |
| 5 | Relaxation of the Forehead | Distortion of the Mouth |
| 6 | Decreased Respiratory Rate | Flinching or Trembling |
| 7 | Upright Sitting Position | Moaning |
| 8 | Laughing out loud | Freezing of the Body |
| 9 | Nodding | Breath Holding |
| 10 | Opening the Fist | Visible Sweating |

**Fig. 4.**
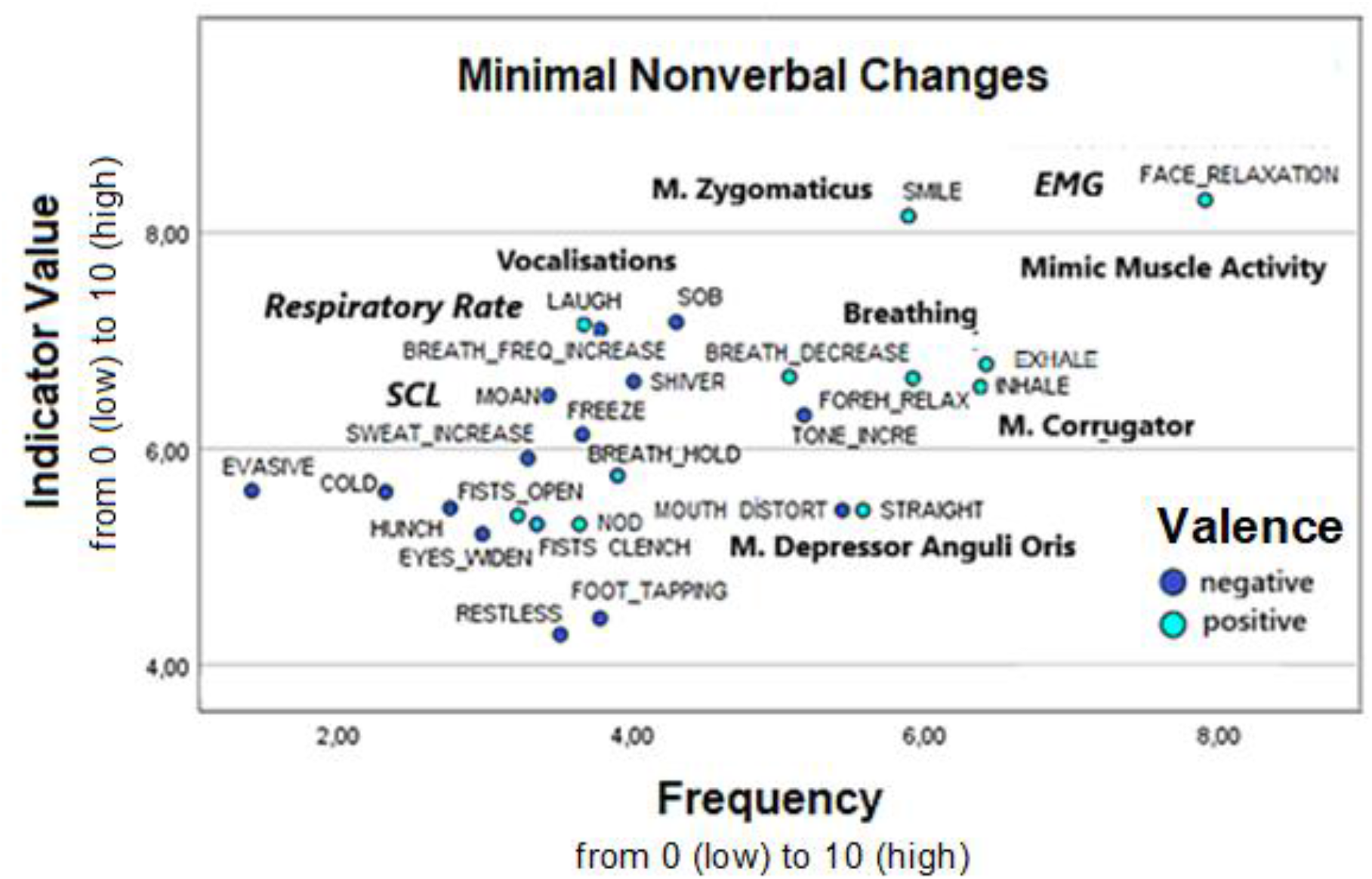
Graphical representation of minimal nonverbal changes (MNCs). Frequency (0-10), indicator value (0-10), and presumed emotional valence (positive, negative) were assessed. Each MNC is represented by a point, with color indicating the presumed emotional valence (negative: blue, positive: green). MNCs with unclear emotional valence (“neutral”) are not shown. Potential physiological correlates are indicated in italics.

## DISCUSSION AND CONCLUSION

The present study provides exploratory insights into how EMDR therapists implement the standard procedure for BLS in routine clinical practice, with a particular focus on the role of MNCs.

### High relevance of MNCs

The majority of therapists reported orienting to MNCs, either as a primary or complementary strategy. Moreover, many therapists reported combining multiple strategies, indicating a flexible and situation-dependent approach rather than strict adherence to a single procedural rule. This pattern suggests that, in clinical practice, MNC-guided approaches may play a central role in decision-making, despite the limited availability of systematic empirical evidence supporting their use. Nevertheless, a minority (15 %) reported relying exclusively counting a fixed number of BLS without adapting to the patient’s individual needs.

### Exploratory analysis of influencing factors

Exploratory analyses of potential influencing factors suggest that variability in procedure implementation may be associated with therapist characteristics, clinical setting, and patient population. However, these findings should be interpreted with caution, given the exploratory nature of the analyses and the absence of correction for multiple testing. Accordingly, the present results should be understood as hypothesis-generating. Further studies using confirmatory designs are required to systematically examine the relationships between therapist-, patient-, and setting-related variables and the implementation of BLS.

#### Impact of therapist characteristics

Exploratory analyses suggest that therapist characteristics may be associated with differences in the implementation of BLS. Descriptively, male therapists, and those with a behavioral orientation tended to report more frequent use of counting strategies. In contrast, female therapists, and those with a psychodynamic orientation more often reported using MNC-based or combined approaches. These patterns may reflect differences in clinical experience and training background. Therapists with a behavioral orientation may rely more strongly on clearly defined procedural rules, whereas clinicians with a psychodynamic orientation may adopt a more flexible, experience-based approach to treatment. Similarly, differences between therapeutic orientations may relate to distinct emphases in training, such as structured intervention strategies in behavioral approaches versus a greater focus on observing internal and nonverbal processes in psychodynamic traditions [32, 33]. Interpretations regarding gender differences [34] should be treated with caution. Although differences in the use of MNCs were observed, the present data do not allow conclusions about underlying mechanisms.

#### Impact of setting characteristics

Differences were also observed between clinical settings. Therapists working in inpatient settings reported more frequent use of counting strategies compared to those working in outpatient settings. One possible explanation is that inpatient populations often present with higher symptom severity and more complex clinical profiles, which may favor the use of more standardized and structured approaches. In addition, reduced emotional expressivity (e.g., due to dissociation or depressive symptoms) may limit the availability of observable MNCs. In contrast, outpatient therapists may have longer-term therapeutic relationships with patients and greater familiarity with individual response patterns, potentially allowing for more flexible and individualized approaches to BLS.

#### Impact of patient characteristics

Therapists treating more complex trauma presentations reported lower willingness to use biofeedback compared to those primarily treating single-event trauma. This finding may reflect differences in clinical complexity. Patients with more complex trauma presentations may be more sensitive to additional external stimuli, such as biofeedback devices, which could be perceived as intrusive or disruptive. In contrast, more standardized approaches may be more readily applicable in less complex cases.

### Awareness of variability and attitudes towards objective alternatives

The observed interindividual variability in the application of BLS may indicate differing levels of certainty among EMDR therapists regarding optimal decision-making during stimulation, particularly with respect to when to continue or interrupt BLS within a set. More objective approaches, such as biofeedback, could represent one potential avenue for standardization. However, the present findings suggest a heterogeneous attitude toward such methods. While a substantial proportion of therapists expressed interest in more explicit criteria and objective indicators, a similarly large group did not endorse these approaches. Notably, therapists working with more complex trauma presentations appeared less inclined toward the use of biofeedback and more likely to rely on individualized clinical judgment. This pattern may reflect differences in perceived clinical demands or concerns about the potential intrusiveness of additional technical procedures.

Overall, these findings highlight a tension between standardization and individualized clinical practice in EMDR. Further research is needed to clarify how different approaches to BLS regulation relate to therapeutic processes and outcomes.

### Frequency, indicator value, and emotional valence of MNCs

A large majority of therapists (98.2%) rated MNCs as important indicators of emotional valence. MNCs were reported to reflect both ongoing negative emotional states and shifts toward more positive valence. MNCs most frequently associated with shifts from negative to positive valence included visible relaxation of the face and body, smiling, laughing, nodding, and deepened breathing. In contrast, MNCs most commonly associated with persistent negative valence included facial tension or distortion, sudden increases in muscle tone, autonomic responses (e.g., sweating, freezing), vocalizations (e.g., sobbing, moaning), and increased or irregular breathing patterns. Therapists reported heterogeneous approaches to handling MNCs. Approximately half indicated no fixed strategy, whereas the other half reported continuing stimulation for a certain period before interrupting. The temporal relationship between MNCs and emotional change was estimated to span both preceding and following phases of valence shifts, with an average latency of a few seconds and substantial variability across respondents.

Overall, these findings are broadly in line with prior research in emotion psychology, suggesting that nonverbal behavior may reflect underlying emotional processes [19–21]. However, given the exploratory and self-reported nature of the data, these observations should be interpreted with caution.

### Implications

The findings indicate substantial variability in how EMDR therapists implement the standard procedure for BLS. This variability suggests a need for clearer operational criteria to guide the timing of BLS interruption, as well as greater consistency across training contexts.

The consistent attribution of high relevance to MNCs by therapists suggests that these cues may play an important role in clinical decision-making. Incorporating structured guidance on the observation and interpretation of MNCs into training programs may support more consistent and transparent application of BLS.

At the same time, reliance on observable MNCs may be limited in patients with low emotional expressivity or subtle physiological responses. In such cases, additional objective indicators may be of interest. Approaches based on physiological measures, such as biofeedback, could represent one possible direction, although their feasibility, acceptability, and clinical utility require systematic investigation.

More broadly, the present findings highlight the need for experimental studies that examine the temporal dynamics of emotional processing during BLS, including the relationship between observable behavior and underlying physiological processes. Such work may contribute to a more precise and empirically grounded definition of optimal set duration.

Finally, future research may also address related clinical challenges, such as the management of prolonged negative processing sequences (e.g., “looping” [3, 35]), for which current approaches vary considerably and lack systematic empirical evaluation.

### Limitations

This study is based on therapists’ self-reports and therefore reflects subjective perceptions rather than directly observed behavior. Response biases, including social desirability and individual judgment tendencies, cannot be excluded. Future studies should complement these findings with observational data, for example through video-based analyses of MNCs during therapy sessions.

Participants were recruited via publicly available directories of EMDR professional associations, which likely introduced self-selection bias. This approach may have resulted in an overrepresentation of particularly engaged and procedure-reflective therapists. Consequently, the observed patterns-such as the high relevance attributed to MNCs and the comparatively lower interest in biofeedback - may not fully generalize to the broader population of EMDR practitioners.

The sample was restricted to therapists from German-speaking countries, limiting generalizability to other cultural and clinical contexts.

The questionnaire was developed for exploratory purposes and provided a predefined set of response options. Additional strategies for determining BLS duration may exist but were not captured. Although the instrument was developed through expert review and iterative revision in line with COSMIN principles [36], formal validation procedures (e.g., content validity indices, pilot testing, and psychometric evaluation) were not conducted. Accordingly, the reliability and validity of the instrument remain to be established in future studies.

For qualitative data, given the exploratory and hypothesis-generating nature of this subsection, no correction for multiple testing was applied. Therefore, reported p-values should be interpreted with caution, as the risk of Type I error is increased and effect sizes may be overestimated.

Finally, ordinal rating-scale data were treated as metric variables, which is common practice but remains methodologically debated [26-27]. This approach may limit the interpretability of the results.

### Conclusion

This survey study demonstrates substantial variability in how certified EMDR therapists implement BLS within the standard procedure. Adaptive strategies based on MNCs were used more frequently than counting approaches. Across respondents, MNCs were consistently rated as highly relevant for guiding BLS interruption, and missing the optimal timing was considered clinically problematic. Specific patterns - particularly changes in facial expression and breathing - emerged as the most promising indicators of emotional valence shifts. Although the findings are exploratory, initial associations between therapist, patient, and setting characteristics and the application of BLS were observed. These results highlight the need for further empirical research to examine the validity and reliability of MNCs as indicators of emotional processing and to establish more precise, evidence-based criteria for determining set duration.

## General

We would like to thank the participating therapists for their interest in our study.

### Ethical Approval

Ethical approval was obtained from the ethics committee of the University Medical Center Rostock.

### Author contributions

V.P. conceived and planned the experiment. M.S. supervised the development of the questionnaire. C.K. performed recruitment and data collection. A.H. contributed to the statistical analyses of the data. M.S., A.H., E.S., and O.W. contributed to the interpretation of the results. V.P. wrote the first draft of the manuscript, and all authors commented on previous versions of the manuscript. English proof-reading was performed by A.H.. All authors read and approved the final manuscript. All authors have read and agreed to the published version of the manuscript.

### Funding

The authors acknowledge that they did not receive funding for this work.

## DATA AVAILABILITY

The raw data of this article is freely available upon request.

## SUPPLEMENTARY MATERIAL

Supplementary Material 1: Questionnaire (*EmdEx*)

